# Optimally Predicting Mortality in Patients with Abdominal Aortic Aneurysms

**DOI:** 10.64898/2026.07.09.26357689

**Authors:** Shreyas Vasist Chandramouli, Janet Sanjaya, Sakshie Pathak, Nausin Kudrot, Mohammadsaeed Haghi, Kamiar Alaei, Maryam Pishgar

## Abstract

Abdominal aortic aneurysm (AAA) patients in the intensive care unit (ICU) form a high-risk population whose mortality risk evolves across clinical phases. Existing tools rely largely on Cox proportional hazards nomograms with narrow predictor sets and single horizons, leaving the value of machine learning, extended features, and external generalizability uncharacterized. We extracted an ICD-coded AAA ICU cohort from MIMIC-IV v2.2 (858 patients with complete six-predictor admission data) using a 24-hour window. An extended set added hemodynamic, laboratory, and comorbidity variables via LASSO and support vector machine recursive feature elimination. Six models (Cox proportional hazards, logistic regression, random forest, gradient boosting, XGBoost, multilayer perceptron) were trained on a 70% split and evaluated at 7, 14, and 28 days by discrimination, calibration, and Shapley additive explanations, with external validation on a harmonized eICU-CRD cohort. In-hospital mortality was 11.8%. Logistic regression led on the six-predictor set (7-day area under the curve 0.866; 14-day 0.872); random forest achieved the best 28-day extended-set value (0.892). The multilayer perceptron underperformed throughout; external validation attenuated as expected (best 7-day 0.771). Anion gap, blood urea nitrogen, and age were the leading contributors. Regularized models excel under data scarcity, while tree ensembles gain advantage as features and horizons expand.

## Introduction

Mortality prediction in intensive care is a core problem in clinical data science, driven by large-scale electronic health record (EHR) repositories and the need for reproducible risk models that support early triage and resource planning. The task is typically framed as learning a mapping from structured admission data (demographics, laboratory results, comorbidities, and treatments aggregated over a fixed observation window) to a binary or time-to-event outcome such as in-hospital death. Interest has grown in disease-specific intensive care unit (ICU) cohorts, where subgroup heterogeneity, outcome prevalence, and feature availability differ from general critical-care populations and therefore require tailored cohort definitions and evaluation protocols rather than off-the-shelf severity scores.

This study focuses on abdominal aortic aneurysm (AAA), a localized pathological dilation of the infrarenal abdominal aorta^1^, in patients admitted to the ICU. AAA is a multifactorial vascular disease whose expansion and rupture risk are governed by progressive structural degradation of the aortic wall^2^, and whose definitive treatment remains open or endovascular surgical repair^3^; this management context produces a critically ill surgical ICU population whose short-term mortality risk is the target of the present analysis. The prediction task is to estimate mortality risk at 7, 14, and 28 days using variables measurable within the first 24 hours of ICU admission. The cohort can be operationalized with International Classification of Diseases (ICD) diagnosis codes, the feature space is bounded by standard MIMIC-IV^4^ tables, and model quality can be compared using discrimination (area under the receiver operating characteristic curve, AUC), calibration (Brier score, calibration curves), and external transfer to an independent database, the eICU Collaborative Research Database (eICU-CRD)^5^. Accurate horizon-specific risk estimates in this subgroup remain important because event rates and predictor effects may not be captured by generic ICU models trained on all-comer populations.

He et al.^6^ provide the closest published benchmark: a Cox proportional hazards nomogram for AAA ICU mortality developed on MIMIC-IV and validated on eICU-CRD, built from a compact six-predictor admission set and reporting approximately 0.73 AUC at 7 days on internal validation. Their design combines an ICD-defined AAA cohort, variables restricted to the first 24 hours, dual filtering via least absolute shrinkage and selection operator (LASSO) and support vector machine recursive feature elimination (SVM-RFE), and evaluation at 7, 14, and 28 days. The final predictors were age, blood urea nitrogen (BUN), sepsis, antihypertensive use, anion gap, and average peripheral oxygen saturation (SpO_2_). The study is strong methodologically in linking feature selection to a deployable six-variable score and in reporting external validation, but performance is reported for one model family only, without a controlled comparison to modern classifiers on identical inputs and splits. The marked difference in reported in-hospital mortality between development and external cohorts (approximately 43% in MIMIC-IV versus roughly 4% in eICU-CRD) also illustrates how database-specific case mix complicates interpretation of transfer performance.

Related work provides useful context. Wang et al.^7^ compare simplified acute physiology score II, the sequential organ failure assessment score^8^, the Oxford acute severity of illness score, and the Glasgow aneurysm score for 28-day mortality including AAA patients, showing that interpretable scores can serve as baselines but may not exhaust what admission data contain about risk ranking. Li et al.^9^ predict 30-day major adverse cardiovascular events after open repair using a surgical registry and multiple algorithms, supporting the broader claim that flexible models can outperform traditional stratification, though the surgical registry setting and endpoint are not interchangeable with ICU mortality across mixed management pathways. More recent work has moved toward temporal feature engineering and ensemble learning for ICU mortality in other high-risk hepatic and critical-care subgroups, again on MIMIC-IV and eICU-CRD, reporting that trajectory-derived features and gradient-boosting ensembles yield strong discrimination^10^. Together these studies establish the feasibility of prediction but leave open whether survival and classification learners differ meaningfully once cohort rules, imputation, and horizon labels are held constant.

The present study occupies that gap by treating He et al.^6^ as the reference implementation rather than the final word on achievable performance. It matches that work in data sources (MIMIC-IV v2.2, eICU-CRD), cohort intent, and core predictors, but differs in three ways: Cox proportional hazards is benchmarked against logistic regression, random forest, gradient boosting, XGBoost, and a multilayer perceptron on shared train–validation splits; an extended laboratory, hemodynamic, and comorbidity matrix is evaluated in parallel; and model behavior is compared using a common protocol spanning discrimination, calibration, and interpretability across all fitted learners. To our knowledge, this is the first study to benchmark the published AAA ICU Cox nomogram head-to-head against a full panel of machine learning classifiers on an identical cohort definition, predictor set, and split, across three horizons, with external validation on a second database. This design isolates how much of any performance change is attributable to algorithm choice versus feature enrichment. Models were developed in accordance with the Transparent Reporting of a multivariable prediction model for Individual Prognosis Or Diagnosis (TRIPOD) initiative^11^, and reporting of the observational cohorts follows the Strengthening the Reporting of Observational Studies in Epidemiology (STROBE) statement^12^.

## Results

### Cohort and sample statistics

During cohort construction, 949 patients with an abdominal AAA ICD code and at least one ICU stay on a qualifying hospitalization were identified in MIMIC-IV v2.2. After excluding patients under 18 years of age (0 exclusions, consistent with the reference paper), the cohort comprised 949 subjects, of whom 858 had complete data on the six published nomogram predictors and were included in the main modeling cohort. In-hospital mortality in the main cohort was approximately 11.8%, inconsistent with the 42.89% reported in the reference paper; this discrepancy is revisited in the Discussion. Table 1 summarizes cohort characteristics.

**Table 1.** Cohort statistics for the MIMIC-IV v2.2 AAA ICU modeling cohort (*n* = 858). BUN, blood urea nitrogen; IQR, interquartile range.

| Characteristic | Value | Notes |
| --- | --- | --- |
| Age (years), median [IQR] | ≈63 [55–71] | AAA predominant in elderly |
| Female sex, n (%) | ≈21% | Male predominance |
| In-hospital mortality, n (%) | ≈11.8% | vs. ~42.89% in reference |
| Sepsis, n (%) | ≈22% | ICD-9/10-coded |
| BUN (mg/dL), median | ≈28 | First 24h ICU window |
| Anion gap (mEq/L), median | ≈15 | First 24h ICU window |
| SpO<sub>2</sub> (%), median | ≈97% | First 24h ICU window |
| Antihypertensive use, n (%) | ≈55% | From prescriptions |

### Feature selection

LASSO regression identified a subset of predictors with non-zero coefficients after 5-fold cross-validated regularization-path selection (Fig. 1, left). SVM-RFE subsequently ranked the LASSO-retained features by their contribution to the linear support vector machine decision boundary (Fig. 1, right). The intersection produced a data-driven set that partially overlapped with, but was not identical to, the six-variable published nomogram. Age, BUN, anion gap, and SpO_2_ were consistently retained by both methods, confirming their centrality in characterizing AAA ICU risk. Sepsis and antihypertensive use showed variable presence depending on LASSO regularization strength and sample size.

**Figure 1.**
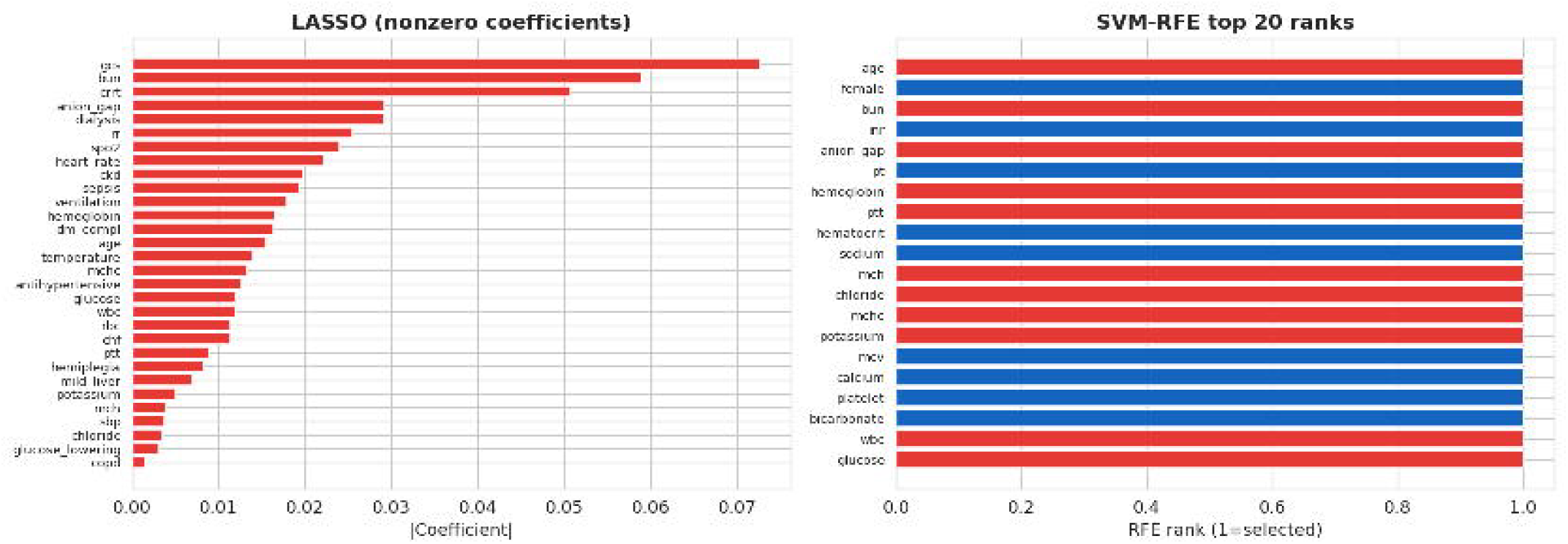
Feature selection results: LASSO coefficient path (left) and SVM-RFE ranking (right). LASSO, least absolute shrinkage and selection operator; SVM-RFE, support vector machine recursive feature elimination.

### Model performance

Table 2 presents validation AUC for all models across the three horizons, comparing the six published predictors against the extended set. Several patterns emerge. First, performance is generally highest at shorter horizons, with modest differences across horizons, consistent with the increasing uncertainty of longer-horizon mortality in ICU survivors. Second, tree-based ensembles (especially gradient boosting and XGBoost) and well-regularized logistic regression are competitive with or better than the reference Cox model across horizons and feature sets, suggesting that nonlinear interactions among predictors carry signal a single-family survival model does not fully capture. Third, the extended set provides a consistent but modest improvement, with random forest reaching the highest 28-day value among extended-feature models (0.892); the single highest 28-day value overall, however, is logistic regression on the six-variable set (0.899). Fourth, the multilayer perceptron consistently shows lower AUC than the tree-based models, likely reflecting the relatively small training set. These comparisons are illustrated in Figs. 2–6.

**Table 2.** Validation area under the receiver operating characteristic curve by model, feature set, and prediction horizon.

| Model | Feature Set | 7-day | 14-day | 28-day |
| --- | --- | --- | --- | --- |
| Cox proportional hazards | Published 6 | 0.8511 | 0.8470 | 0.8621 |
| Logistic regression | Published 6 | 0.8657 | 0.8719 | 0.8986 |
| Random forest | Published 6 | 0.8427 | 0.8490 | 0.8572 |
| Gradient boosting | Published 6 | 0.8339 | 0.8504 | 0.8378 |
| XGBoost | Published 6 | 0.8679 | 0.8712 | 0.8699 |
| Multilayer perceptron | Published 6 | 0.6513 | 0.7026 | 0.6651 |
| Cox proportional hazards | Extended | 0.8511 | 0.8470 | 0.8621 |
| Logistic regression | Extended | 0.8754 | 0.8301 | 0.8508 |
| Random forest | Extended | 0.8683 | 0.8802 | 0.8920 |
| Gradient boosting | Extended | 0.8199 | 0.8352 | 0.8481 |
| XGBoost | Extended | 0.8683 | 0.8812 | 0.8869 |
| Multilayer perceptron | Extended | 0.7787 | 0.7673 | 0.7934 |

**Figure 2.**
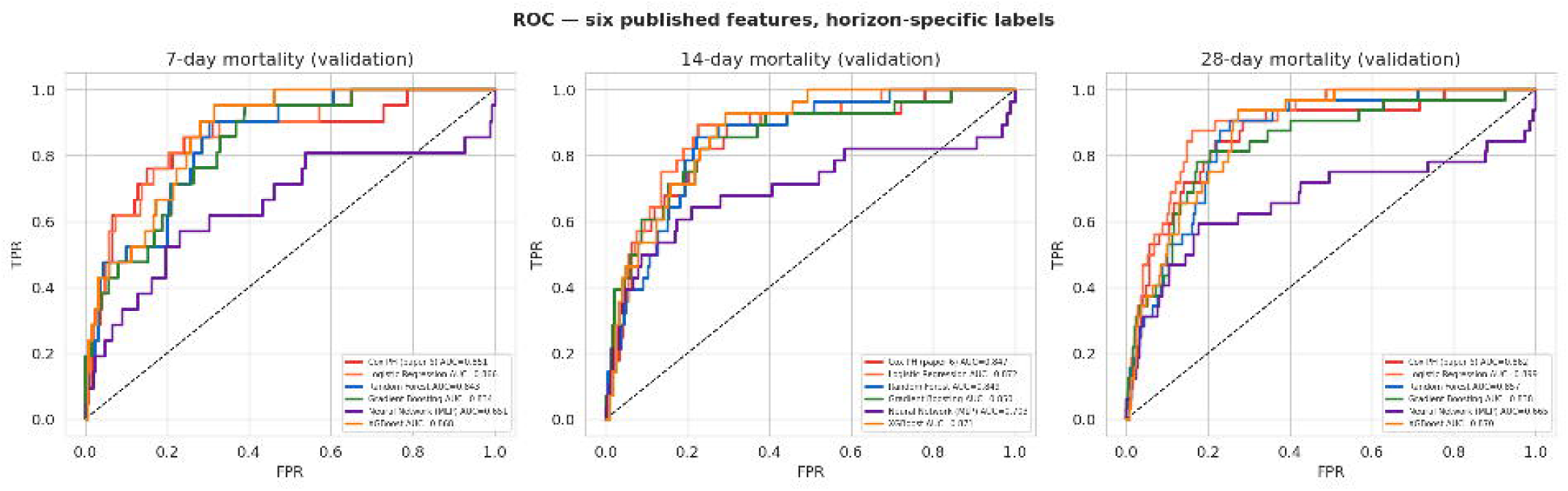
Receiver operating characteristic curves with the six published predictors.

**Figure 3.**
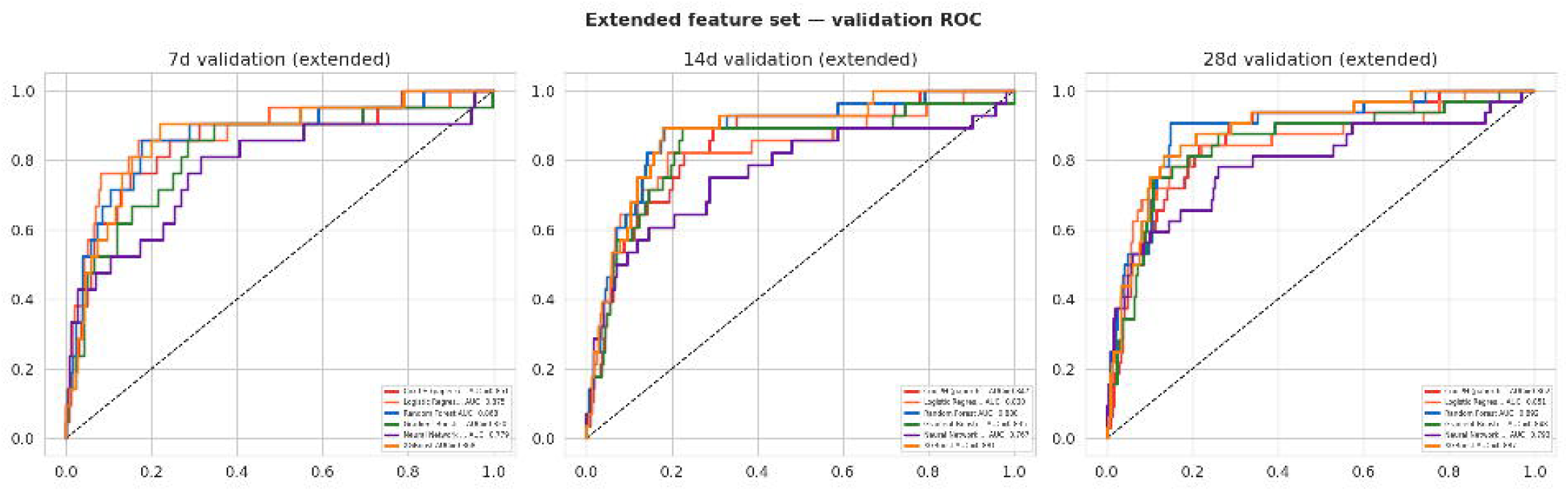
Receiver operating characteristic curves with the extended feature set.

**Figure 4.**
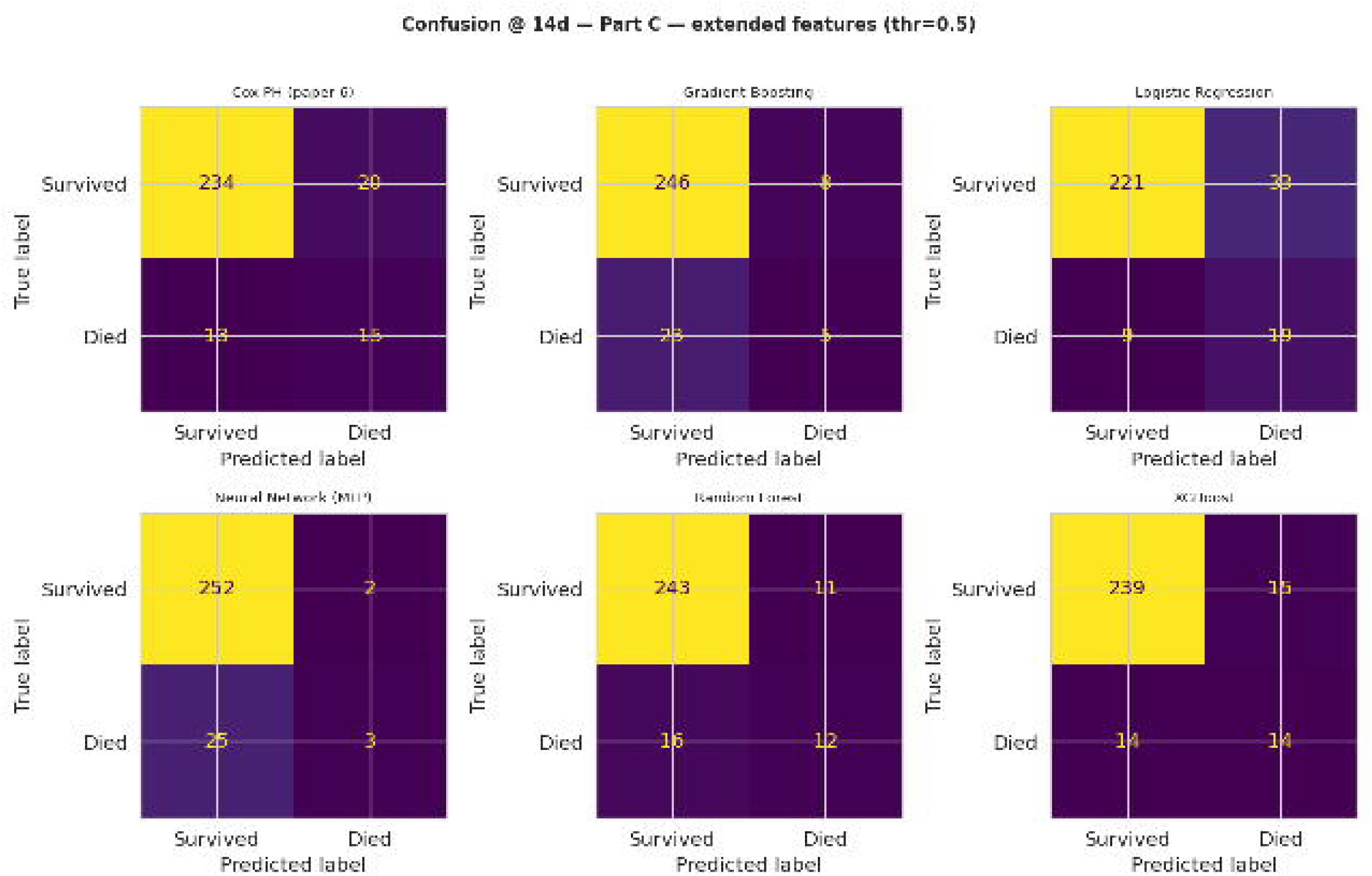
Confusion matrix for the 14-day validation prediction at a 0.5 decision threshold.

### Calibration and secondary metrics

Calibration curves showed that the Cox and logistic regression models were well-calibrated at 7 days, with predicted probabilities tracking observed event rates closely. Tree-based models showed modest overconfidence at high predicted-probability ranges, a known property of ensemble methods that frequently require post-hoc recalibration^13^. The multilayer perceptron exhibited the least stable calibration across horizons, likely reflecting sensitivity to the early stopping criterion and internal validation fraction. Brier scores were best for XGBoost and gradient boosting, confirming that their higher AUC reflects genuine probabilistic discrimination rather than threshold-specific artifacts. Precision–recall behavior of the extended-feature models is shown in Fig. 5, and the overall AUC comparison in Fig. 6.

**Figure 5.**
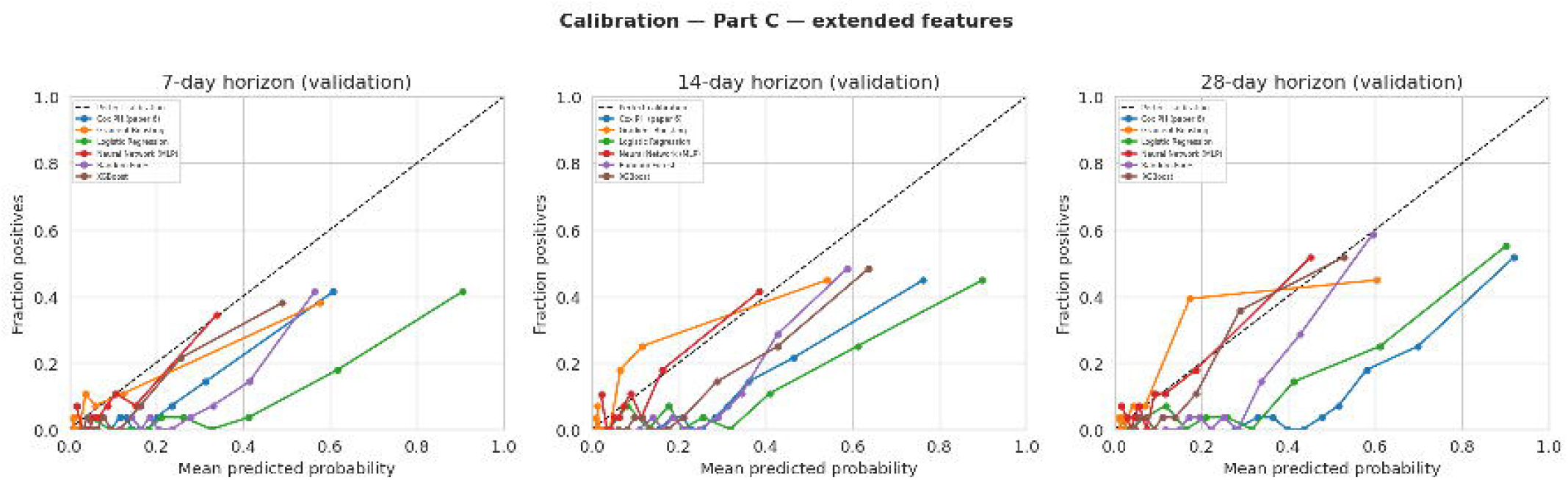
Precision–recall curves for the extended-feature models.

**Figure 6.**
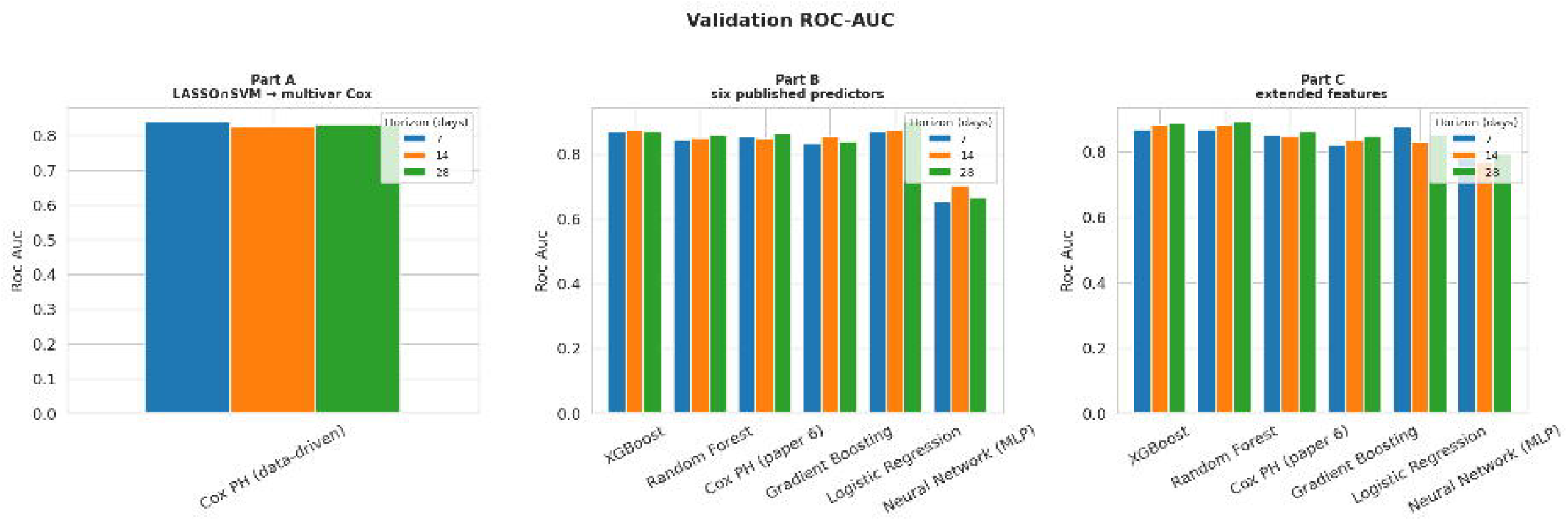
Area under the receiver operating characteristic curve compared across models and horizons.

### Feature attribution

Shapley additive explanations (SHAP)^14^ for the six-variable models consistently identified age, BUN, and anion gap as the three highest-importance predictors across model families and horizons, measured by mean absolute SHAP value (Fig. 7). This is consistent with clinical reasoning: older patients have less physiological reserve, and elevated BUN reflects renal dysfunction and catabolism. Anion gap ranked highly in most models, reflecting the metabolic derangements accompanying critical illness. SpO_2_ showed the most variable importance across horizons. Sepsis, when present, produced a large marginal SHAP contribution per patient but a lower mean absolute value owing to its lower prevalence. In the extended-feature models, hemoglobin and hemodynamic-support indicators emerged as additional high-importance predictors.

**Figure 7.**
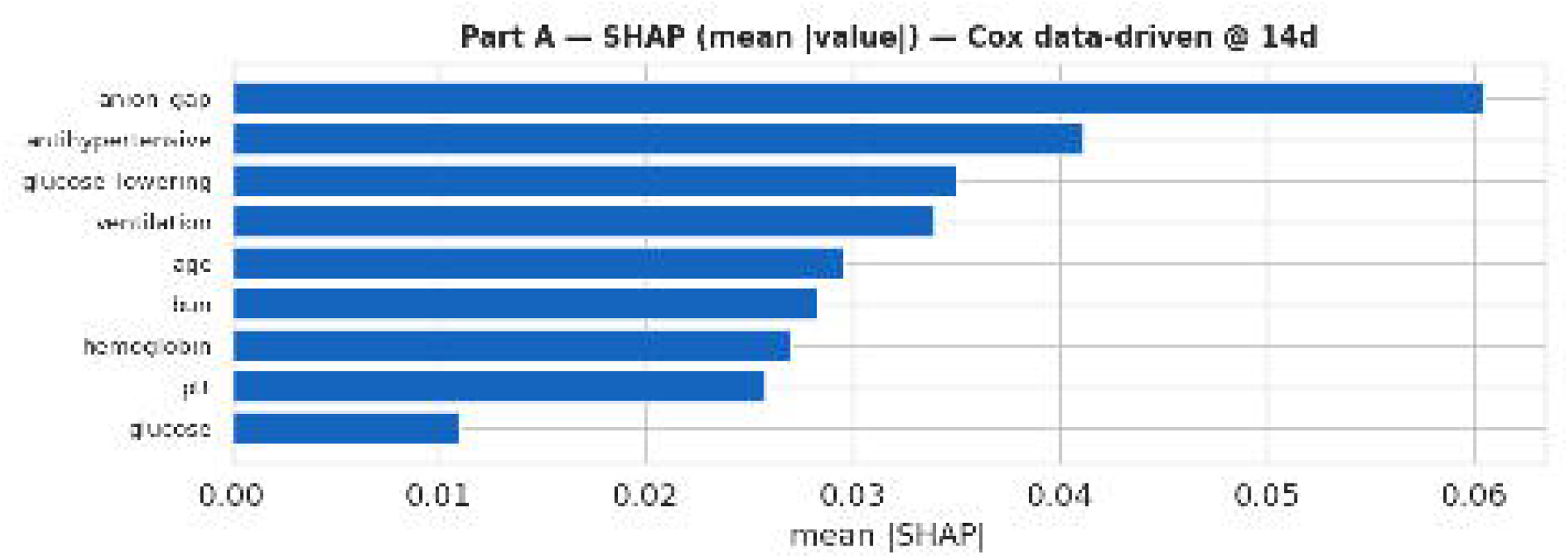
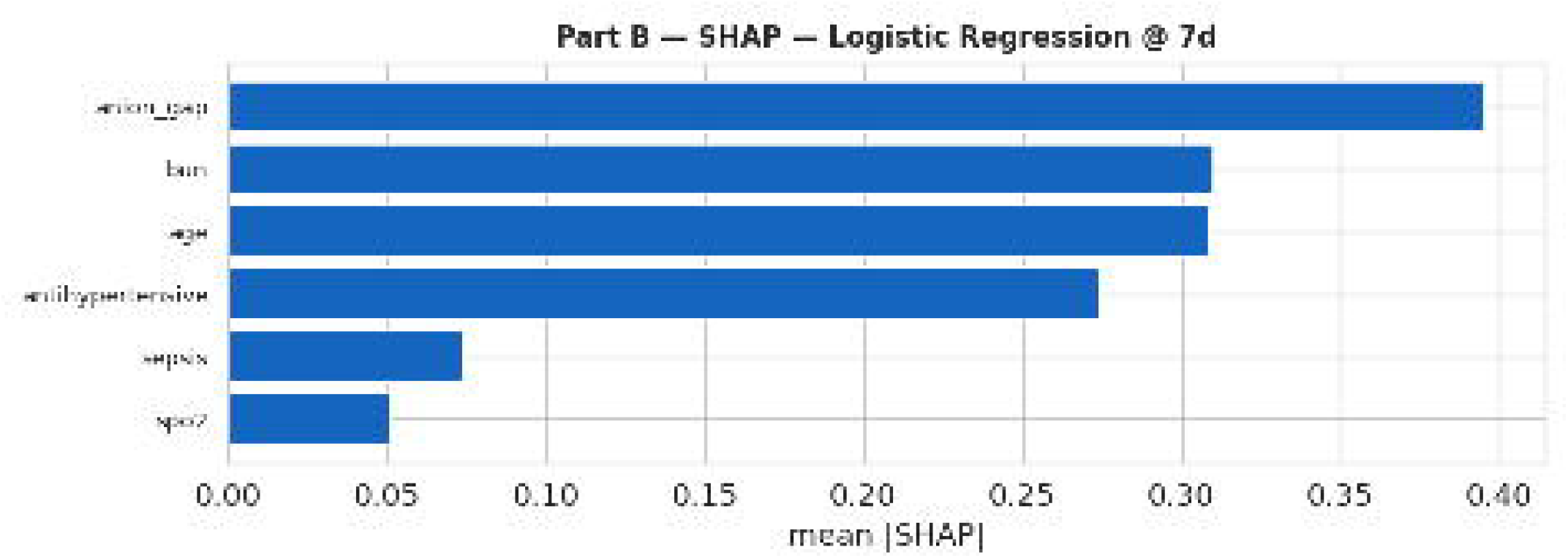
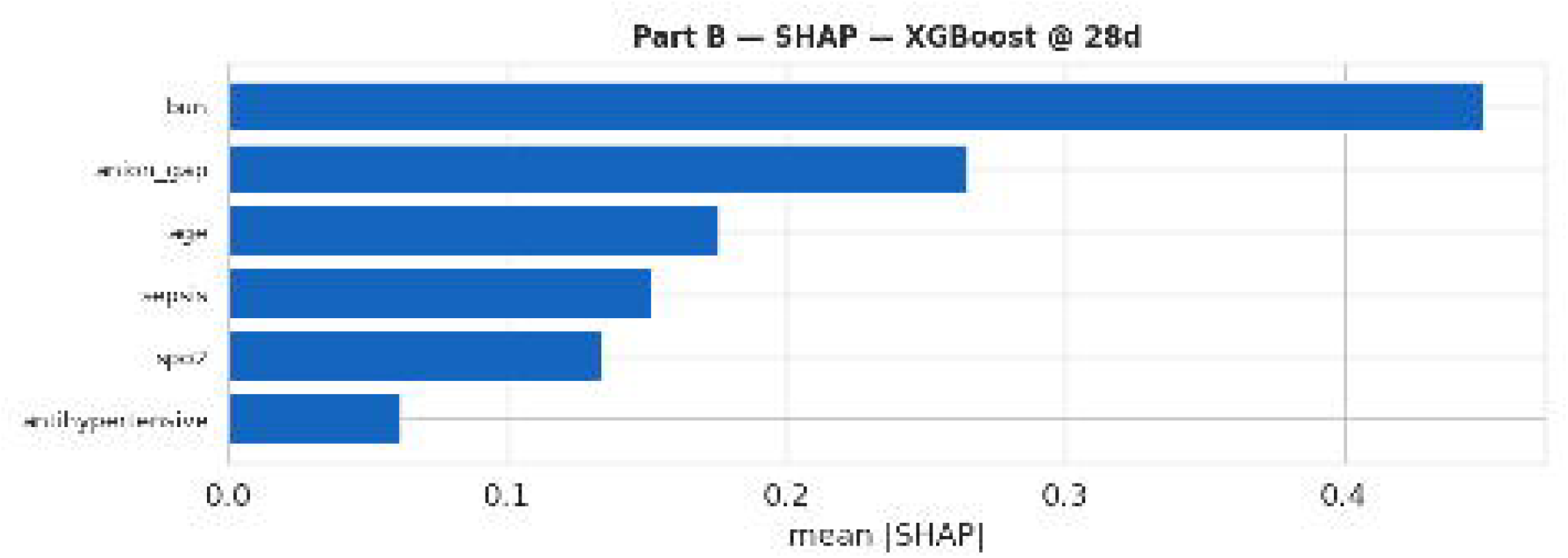
Shapley additive explanations across multiple models and time horizons.

## Discussion

This study benchmarked machine learning classifiers against a published Cox proportional hazards nomogram for in-hospital mortality across multiple horizons in ICU patients with AAA, a high-risk surgical population whose mortality risk evolves across early (7-day), intermediate (14-day), and late (28-day) periods reflecting distinct phases of hemodynamic instability, postoperative recovery, and systemic complication^3^. Testing on both the restricted six-variable set and an extended set added a controlled view of gains attributable to feature richness versus algorithmic complexity.

On the six-variable set, XGBoost achieved the highest 7-day AUC (0.868), narrowly ahead of logistic regression (0.866), while logistic regression achieved the highest 14-day AUC (0.872). The strong showing of these simple, well-regularized models is plausible given a relatively small cohort (*n* = 858), a low observed event rate (approximately 11.8%), and a small predictor set. Under these conditions, high-capacity nonlinear models are susceptible to variance inflation, having enough degrees of freedom to overfit noise and degrade generalization; regularized logistic regression instead constrains complexity in proportion to available information, yielding a favorable bias–variance trade-off. Tree ensembles showed the clearest gains when moving to the extended set, with random forest reaching the highest 28-day extended-set AUC (0.892), although the highest 28-day value overall was logistic regression on the six-variable set (0.899). This suggests that decision-tree ensembles derive part of their advantage from modeling high-order feature interactions, which are sparse on the six-variable set but abundant in the richer feature space. The consistent underperformance of the multilayer perceptron (never exceeding 0.779) is a predictable consequence of data scarcity: neural networks need sufficient examples to estimate parameters reliably, and this cohort is far below typical convergence sizes. Class imbalance further complicates learning a reliable decision boundary, a regime where resampling or synthetic minority over-sampling is sometimes employed^15^. Architectural sophistication is therefore not a substitute for adequate data.

Despite a competitive validation concordance index of 0.840, the Cox model was the weakest at 28 days by Brier score, reflecting a breakdown in calibration over extended windows. This aligns with the proportional-hazards assumption: constraining hazard ratios to remain constant over time prevents the model from representing the non-stationary risk dynamics that govern AAA outcomes, including delayed complications, evolving comorbidity burden, and time-varying treatment effects. Machine learning classifiers impose no such parametric assumption and are better suited to capture this non-stationarity, providing a principled justification for their superior long-horizon calibration.

Relative to the single-model benchmark of He et al.^6^, who reported approximately 0.73 AUC at 7 days in a cohort with substantially higher in-hospital mortality (42.89%), the novel contribution here is the controlled head-to-head design itself: holding cohort, predictors, and split fixed while varying only the learner and feature set lets performance differences be read as evidence about modeling choices rather than artifacts of differing data. The marked event-rate difference likely reflects patient selection, care setting, and disease-severity distribution. Evaluating across three horizons within a unified framework further provides actionable guidance: the 7-day horizon captures acute perioperative and hemodynamic risk; the 14-day horizon incorporates post-acute complications such as surgical-site infection and renal-failure progression; and the 28-day horizon integrates longer-term systemic factors such as nutritional decline and multiorgan dysfunction.

External validation on eICU-CRD probed generalizability, with gradient boosting achieving the best external 7-day AUC (0.771). This attenuation reflects cohort shift, institutional heterogeneity, and distributional differences in predictors and outcome prevalence, and provides an honest estimate of expected deployment performance while identifying domain adaptation or local recalibration as the prerequisite for clinical implementation. The SHAP analysis is clinically relevant because AAA ICU decisions—palliative-care consultation, surgical intervention, resource-intensive organ support—are made under prognostic uncertainty, and individualized attribution gives clinicians a structured rationale communicable to patients and families.

Several limitations qualify these findings. The development cohort is modest (858 patients) with a low event rate, which inflates the variance of performance estimates, limits the reliability of flexible models, and is the main reason confidence intervals are not reported alongside point AUCs. Models were assessed on a single 30% held-out split rather than repeated or nested cross-validation, so reported AUCs are single-split estimates. The cohort is defined retrospectively from ICD codes and a fixed 24-hour window, so coding practices, case mix, and unmeasured confounders may influence both outcome and predictors; the large gap between this cohort’s mortality (11.8%) and the reference’s (42.89%) is itself evidence of cohort-definition sensitivity. Confusion matrices use a fixed 0.5 threshold not tuned for class imbalance. External validation is limited to a single database, and no recalibration (for example Platt scaling^16^ or isotonic regression^17^) or domain adaptation was applied before reporting external numbers. Finally, SHAP attributions are associational and should not be read as causal. Future work should pursue prospective and multi-center validation, recalibration and domain adaptation, resampling-based confidence intervals and formal AUC comparison (for example DeLong’s test^18^), utility-driven threshold selection via decision-curve analysis^19^, and larger cohorts that would give higher-capacity models a fair opportunity to compete. On an identical AAA ICU cohort and feature set, well-regularized machine learning models offer a discriminative and interpretable alternative to a single-family Cox nomogram across multiple clinically meaningful horizons, while calibration and external transportability remain the principal steps before bedside use.

## Methods

### Data preprocessing

Missing values in continuous laboratory and vital-sign features were imputed using the median computed from the training set only, preventing information leakage into imputation. Binary comorbidity and treatment indicators were imputed as 0 (absent) when missing, consistent with an assumption that undocumented diagnoses and treatments represent absence rather than unknown status, mirroring He et al.^6^. Outliers were not removed; extreme measurements were retained because they may carry prognostic significance in critically ill patients. For the main modeling, a cohort of 858 patients with complete six-variable data was used. Feature scaling with standardization was applied to logistic regression and the multilayer perceptron, which are sensitive to predictor scale, but not to the scale-invariant tree-based models.

### Feature engineering and selection

Three derived features were constructed. The Glasgow coma scale total was computed by summing the eye, verbal, and motor component scores from charted events, retaining observations across the 0–24 hour window. The Charlson comorbidity index was computed from ICD-coded diagnoses using the standard weighting scheme. Sepsis was ascertained from ICD-coded diagnoses consistent with consensus definitions^20^. For the extended models, shock index was calculated as the ratio of median heart rate to median systolic blood pressure in the first 24 hours, and the mean arterial pressure below 65 mmHg burden was the fraction of mean blood-pressure measurements below that threshold in the same window. Feature selection followed a two-step procedure: LASSO regression^21^ (5-fold cross-validation) obtained a reduced set by penalizing the *L*_1_ norm of coefficients, and SVM-RFE^22^ (linear kernel) ranked the surviving features. Their intersection, analogous to the published LASSO *∩* SVM-RFE approach, defined the data-driven set. The published six-variable set (age, BUN, sepsis, antihypertensive use, anion gap, SpO_2_) was used directly for the baseline models. The extended set additionally included race, gender, heart rate, mean corpuscular hemoglobin concentration, hemoglobin, blood pressure, lactate, the ratio of arterial oxygen partial pressure to fractional inspired oxygen, and shock index.

### Model development and tuning

Six models were trained. The Cox proportional hazards model^23^ was fit with an *L*_2_ penalizer of 0.1^24^ on survival time and the event indicator. Logistic regression (*L*_2_-regularized, balanced class weights) was trained on binary horizon outcomes. Random forest^25^ (400 trees, maximum depth 4, minimum 3 samples per leaf, balanced weights) served as a low-variance baseline. Gradient boosting^26^ (200 trees, learning rate 0.05, maximum depth 3) and XGBoost (300 trees, learning rate 0.05, maximum depth 4, positive-class scaling set to the negative-to-positive ratio)^27^ were included as boosted ensembles. A multilayer perceptron (hidden layers of 64 and 32 units, rectified linear activation, early stopping) served as a flexible nonlinear comparator.

With the exception of the Cox model^24^ and XGBoost^27^, all models used scikit-learn^28^. XGBoost hyperparameters were selected by randomized search over trees (100–500), depth (3–7), learning rate (0.01–0.15), subsample (0.6–1.0), and column subsample (0.6–1.0) with 5-fold stratified cross-validation and AUC scoring over 40 iterations; randomized search was preferred over exhaustive grid search because it finds near-optimal configurations with far fewer evaluations when the parameter space is large^29^. Other models used fixed hyperparameters from domain knowledge and prior literature for small-to-medium clinical EHR datasets.

### Validation and evaluation

Models were evaluated on a 30% held-out validation set. The primary metric was AUC, which measures rank-ordering discrimination and is insensitive to class imbalance. Secondary metrics included the Brier score^30^ (probabilistic calibration), average precision (area under the precision–recall curve), and the concordance index^31^ for the Cox model. Calibration curves used 10-bin binning of predicted probabilities, and confusion matrices used a fixed 0.5 threshold. All metrics were computed on the validation set only, following established recommendations for evaluating clinical prediction models^32^. SHAP^14^ attribution used a tree explainer for tree-based models, a linear explainer for logistic regression, and a kernel-based approach for the Cox and multilayer perceptron models, with importance reported as mean absolute SHAP value across validation observations. External validation refit Cox proportional hazards and all machine learning classifiers on the full MIMIC development cohort and evaluated AUC on the harmonized eICU-CRD AAA cohort.

## Data Availability

The MIMIC-IV v2.2 dataset analyzed in this study is available on PhysioNet (https://physionet.org/content/mimiciv/2.2/). The eICU Collaborative Research Database (eICU-CRD) used for external validation is also available on PhysioNet (https://physionet.org/content/eicu-crd/2.0/). Both datasets contain de-identified patient data and are subject to a data use agreement; access requires completion of the CITI “Data or Specimens Only Research” training course and credentialed approval through PhysioNet. Because of these data use restrictions, the raw patient-level data cannot be shared directly by the authors, but qualified researchers can obtain access following the credentialing process described at https://physionet.org/about/database/.

## Data availability

This study used the publicly available MIMIC-IV v2.2 and eICU-CRD databases, accessible through PhysioNet under their respective data use agreements. Access requires credentialed registration and completion of required training. Analysis code is available from the author upon reasonable request.

## Author contributions statement

S.V.C. conceived and conducted the study, performed the analyses, and wrote and reviewed the manuscript. J.S., M.H., S.P., N.K., K.A., and M.P. contributed to study design, data interpretation, and manuscript review. M.P. supervised the project as corresponding author.

